# Reduced risk of SARS-CoV-2 infection among household contacts with recent vaccination and past COVID-19 infection: results from two multi-site case-ascertained household transmission studies

**DOI:** 10.1101/2023.10.20.23297317

**Authors:** Melissa A. Rolfes, H. Keipp Talbot, Kerry Grace Morrissey, Melissa S. Stockwell, Yvonne Maldonado, Huong Q. McLean, Karen Lutrick, Natalie M. Bowman, Suchitra Rao, Hector. S. Izurieta, Yuwei Zhu, James Chappell, Steph Battan-Wraith, Lori S. Merrill, Son McClaren, Ellen Sano, Joshua G. Petrie, Jessica Biddle, Sheroi Johnson, Philip Salvatore, Sarah E. Smith-Jeffcoat, Edwin J. Asturias, Jessica T. Lin, Katherine D. Ellingson, Edward A. Belongia, Vanessa Olivo, Alexandra M. Mellis, Carlos G. Grijalva, Respiratory Virus Transmission Network Study Group

## Abstract

**Background:** COVID-19 vaccines reduce the risk of severe disease, but it is less clear what effect vaccines have on reducing the risk of infection in high contact settings like households, alone or in combination with prior infection.

**Methods:** Households with an individual who tested positive for SARS-CoV-2 during Sep 2021–May 2023 were screened nationwide and at 7 sentinel sites and enrolled if the index case’s illness onset was ≤6 days prior. Household members had daily self-collected nasal swabs tested by RT-PCR for SARS-CoV-2. COVID-19 vaccination status was assessed by plausible self-report (with date) or vaccination records. Prior infection was assessed by self-reported prior testing and by anti-nucleocapsid antibodies presence at enrollment. The effects of prior immunity, including vaccination, prior infection, or hybrid immunity (both vaccination and prior infection) on SARS-CoV-2 infection risk among household contacts were assessed by robust, clustered multivariable Poisson regression.

**Findings:** There were 1,532 contacts from 905 households included in this analysis. Of these, 67% were enrolled May–November 2022, when Omicron BA.4/5 predominated. Most contacts (89%) had some immunity to SARS-CoV-2 at the time of household exposure: 8% had immunity from prior infection alone, 51% from vaccination alone, and 29% had hybrid immunity. Sixty percent of contacts tested SARS-CoV-2-positive during follow-up. The risk of SARS-CoV-2 infection was not significantly reduced by vaccination but was reduced among those with prior infection considering such immunity separately (adjusted relative risk 0.83; 95% confidence interval: 0.77, 0.90); however, when accounting for both sources of immunity, only contacts with vaccination and prior infection had significantly reduced risk of infection (aRR: 0.81, 95% CI: 0.70, 0.93). The risk of infection was lower when the last immunizing event (vaccination or infection) occurred ≤6 months before COVID-19 affected the household (aRR: 0.69, 95% CI: 0.57, 0.83).

**Interpretation:** Immunity from COVID-19 vaccination and prior infection was synergistic in protecting household contacts from SARS-CoV-2 infection. These data support COVID-19 vaccination, even for those who have been previously infected.

**Research in context:** *Evidence before this study:* We searched PubMed using the terms (“hybrid immunity” or “natural immunity”) AND (“SARS-CoV-2” or COVID*) in October of 2023 to identify previous research into the role of hybrid immunity (defined as immunity from prior infection and vaccination) in susceptibility to SARS-CoV-2 infections. We reviewed 512 articles for estimates of the association between hybrid immunity and susceptibility to illness, infection, or reinfection in humans. Multiple previous meta-analyses were identified, including a meta-regression from 2023 finding that hybrid immunity was associated with 61% reduction in risk of infection compared to immune-naïve individuals 6 months after the immunizing event. The estimates included in this meta-regression were all published before June of 2022, prior to the widespread circulation of Omicron BA.4, BA.5, or recombinant lineages, and none reported on the risk of infection in a setting of household exposures.

*Added value of the study:* In a pair of multi-site case-ascertained household transmission investigations with the majority of enrollments occurring during the Omicron BA4/5 predominant periods, the risk of infection among household contacts of a person with SARS-CoV-2 infection was high. In a study design with systematic, daily testing of household contacts regardless of symptoms, serological verification of prior infection, and vaccine verification, the primary result of analyses of infection risk among household contacts was that this risk was lowest among those with hybrid immunity. The estimate of the magnitude of this protection was lower than in previous reports of protection in other settings.

*Implications of all the available evidence:* The risk of SARS-CoV-2 infection among household contacts was lowest among those with hybrid immunity, compared to those with no previous immunity, vaccination alone, or previous infection alone. These findings underscore the importance of staying updated with COVID-19 vaccinations, even for individuals with prior infection.

## Introduction

COVID-19 vaccines reduce the risk of symptomatic infection, hospitalization, and death from SARS-CoV-2 infection.^1, 2^ However, vaccine-induced protection wanes over time,^3^ and it is less clear whether vaccination protects against mild, non-medically attended SARS-CoV-2 infections, which may comprise more than two-thirds of cases.^4^ Immunity derived from surviving a prior SARS-CoV-2 infection also reduces the risk of subsequent infection and severe or fatal disease.^2, 5, 6^ As the COVID-19 epidemic continues, hybrid immunity, derived from vaccination and prior infection, is becoming widespread.^2, 7^ While vaccine-induced and infection-induced immunity may behave synergistically,^8^ few studies have characterized the risk of infection by hybrid immunity during the Omicron era or in other settings with intense exposures and high risk of transmission. ^9–16^

Households account for a substantial proportion of SARS-CoV-2 transmission, with risk of transmission in this setting up to three times higher than in other settings.^17^ Understanding protection afforded by COVID-19 vaccination, prior SARS-CoV-2 infection, or both in these high-risk settings is important. To this end, two large multi-site case-ascertained household transmission studies were conducted in the United States to estimate the infection risk among household contacts exposed to SARS-CoV-2 and characterize whether COVID-19 vaccination and/or prior infection mitigates that risk.

## Methods

### Study population

Two case-ascertained household transmission studies were conducted across the United States, during September 2021 to May 2023. In one study (the sentinel approach), seven sentinel sites (in Arizona, California, Colorado, New York, North Carolina, Tennessee, and Wisconsin) identified and recruited individuals who recently tested positive for SARS-CoV-2 at academic outpatient medical centers, health systems, or through ongoing surveillance or public health registries. In the second study (the national approach), individuals who tested positive for SARS-CoV-2 at outpatient clinics, retail pharmacies, or through at-home testing services, or who self-reported a positive result through participatory surveillance systems, were identified and invited to participate in the study. For the national approach, potential participants were recruited in one of two ways: a) contacted by email or phone by affiliated service providers, or b) information provided about the study with a corresponding link posted on research message boards, online. For both approaches, individuals who tested positive for SARS-CoV-2 (index cases) were enrolled if: they were the first individual in the household with evidence of infection in the past week (primary case); enrollment occurred within 6 days of index illness onset or first positive test if the index case was asymptomatic; and the index case planned to live in the household during the study follow-up period. Household contacts were screened and enrolled if they routinely slept in the same household as the index case and planned to do so during the study follow-up period (defined as sleeping in the same household at least half the nights in the past month, and at least once since the date of symptom onset in the index case). Households were eligible for inclusion if at least two-thirds of all possible household contacts enrolled in the study; reasons for ineligibility prior to enrollment are given in Supplemental Figure 1 for the national approach, and Supplemental Figure 2 for the sentinel approach. The target enrollment number was 1,400 households, to provide >80% power to detect a 30% relative difference in infection risk between two exposure groups (e.g., vaccinated or unvaccinated) if the baseline risk of infection was 15% or more and assuming similar within-household correlation as reported in prior household transmission studies.^18^ Study activities were reviewed and approved by Vanderbilt University and sites participating (with Vanderbilt University serving as the Institutional Review Board) in the sentinel approach and the Westat Institutional Review Board in the national approach (see 45 C.F.R. part 46.114; 21 C.F.R. part 56.114).

At enrollment, index cases and household contacts self-reported demographic characteristics, vaccination status, and history of prior SARS-CoV-2 infection (history of positive test(s) and year and month of most recent positive test) and self-collected a blood specimen. All participants retrospectively reported symptoms experienced between index case illness onset and study enrollment. Participants completed daily symptom and medication diaries and self-collected (or for young children, adult-collected) nasal swabs (in the sentinel approach) or saliva (in the national approach) prospectively for ten days. Study staff verified vaccination status through state immunization registries, medical chart reviews, calls to vaccination providers, and issued vaccination cards.

### Laboratory methods

Participants collected at-home blood using Asanté dried blood specimen collection strips or Neoteryx Mitra Cartridge devices (in the sentinel approach), or Tasso+ Kits (in the national approach) at enrollment. In the sentinel approach, eluted specimens were tested for evidence of anti-nucleocapsid antibodies using the Thermo ProcartaPlex assay on the Luminex MAGPIX platform at Vanderbilt University Medical Center. Specimen median fluorescence intensity (MFI) values were normalized to kit-specific low-control MFI values, and ratios were used to individually classify nucleocapsid antibodies as absent (<1), present (>1.3), or indeterminate (1-1.3) as specified in manufacturer’s protocols. In the national approach, eluted specimens were tested using the COBAS Elecsys anti-SARS-CoV-2 assay at CoreMedica, and cutoff index (COI) values <1 were considered negative.

Nasal specimens were tested by real-time quantitative reverse-transcriptase polymerase chain reaction (qRT-PCR) using the Panther Fusion Hologic system at Vanderbilt University Medical Center. Saliva specimens were tested using the Infinity BiologiX TaqPath SARS-CoV-2 Assay (EUA: 137776).

### Statistical analysis

For analysis, a COVID-19 vaccination dose was considered verified if there was documentation in the examined records or plausible if it was self-reported with a date (at least the month and year were reported) and the location of vaccination or the manufacturer of the vaccine were reported; if not plausible or verified, the vaccination dose was not counted. Participants with 2 or more verified or plausibly reported doses of COVID-19 vaccine received 14 or more days prior to the index case’s illness onset/clinical test date were considered vaccinated; participants with 0 or 1 verified or plausibly reported dose received 14 or more days prior to study enrollment were considered unvaccinated for initial analyses. Participants were considered age-ineligible for vaccination if they were not eligible for vaccination based on their age 14 days before the household was enrolled.^19^ The number and date of last verified or plausibly reported COVID-19 vaccine doses were calculated. The valency of each dose (monovalent or bivalent; bivalent vaccines were formulated based on the SARS-CoV-2 ancestral strain and a BA.4/BA.5 Omicron strain) was also captured by self-report and in vaccine records.

Participants who had anti-nucleocapsid antibodies detected at enrollment or participants who reported a prior positive test for SARS-CoV-2 along with the year of the last positive test were considered to have prior SARS-CoV-2 infection. The date of prior SARS-CoV-2 infection was assumed to be the date provided through self-report; the date of prior SARS-CoV-2 infection was unknown for participants who did not report a prior positive test but had anti-nucleocapsid antibodies detected. Prior positive tests reported by participants that occurred in the same month as enrollment may reflect a current, rather than prior, infection and thus were not considered when defining prior SARS-CoV-2 infection.

Participants were considered to have hybrid immunity if they had received at least 2 doses of COVID-19 vaccine and had any prior SARS-CoV-2 infection. The number of months since the last immunizing event was calculated as the absolute difference in months from enrollment to the most recent COVID-19 vaccine dose or last positive SARS-CoV-2 test. Hybrid immunity, with monovalent alone or including bivalent COVID-19 vaccination, was also examined.

Analysis excluded households with two or more likely primary cases and household contacts who did not collect their first respiratory specimen within 7 days of the index case’s illness onset, withdrew from the study, had <2 respiratory specimens tested, or did not respond to questions on COVID-19 vaccination and prior SARS-CoV-2 testing.

Among household contacts, the risk of RT-PCR-confirmed SARS-CoV-2 infection was estimated considering COVID-19 vaccination, prior SARS-CoV-2 infection, and hybrid immunity status. Statistical comparisons of risk were conducted using generalized estimating equations, with clustering at the household level and assuming an exchangeable correlation structure and a Poisson framework. Unadjusted models were examined along with models adjusted for the recruitment approach for the household (national or sentinel), the age of the household contact (<5, 5–11, 12–17, 18–49, ≥50 years), household density (number of people/number of bedrooms), and month of enrollment (as a continuous variable from September 2021). Planned secondary analyses included stratification by recency of immunizing events (<6 months vs. ≥6 months), the number and recency of vaccine doses received, and prior infection.

Several planned sensitivity analyses were conducted to examine the robustness of the findings (**Supplemental Table 1**). To reduce potential misclassification of COVID-19 vaccination, a sensitivity analysis was conducted with vaccination defined by vaccine records verification only and participants who self-reported doses that were not verified by records were excluded. To reduce potential misclassification of prior SARS-CoV-2 infection status, three sensitivity analyses were conducted: 1) restricted to household contacts with complete antibody results and defining prior infection only by the detection of anti-nucleocapsid antibodies; 2) defining prior SARS-CoV-2 infection as detection of anti-nucleocapsid antibodies or self-reported prior positive test with month and year of test reported; and 3) using the definition of prior SARS-CoV-2 infection from the main analysis but excluding households that had any participant that reported a positive SARS-CoV-2 test within the three months before enrollment. Another sensitivity analysis was conducted with hybrid immunity restricted to participants with completed antibody testing and verified vaccine doses using the following definitions: participants with hybrid immunity were those with anti-nucleocapsid antibodies and ≥2 verified COVID-19 vaccine doses, only vaccination immunity were those with ≥2 verified vaccination doses and no detected anti-nucleocapsid antibodies, only prior SARS-CoV-2 infection were those with detected anti-nucleocapsid antibodies and <2 verified doses of COVID-19 vaccination, and no immunity were those with <2 verified doses of COVID-19 vaccine and no detected anti-nucleocapsid antibodies. Another analysis was conducted with hybrid immunity where vaccination with ≥1 dose was sufficient. To understand whether model results were robust by study recruitment approach (including differences in laboratory methods), the fully adjusted models were run on data from the sentinel and national approaches separately. Additional analyses were conducted examining hybrid immunity separately by whether participants had received bivalent COVID-19 vaccination.

Data management and statistical analyses were conducted in R Statistical Software (version 4.2.3, R Core Team, 2023).

## Results

### Study population

During September 2021–May 2023, 14814 index cases with laboratory confirmed SARS-CoV-2 infection were screened and 1,482 households with 3,884 individuals, including 2,402 household contacts, were enrolled in the case-ascertained transmission studies. After excluding 870 household contacts (114 withdrew from the study, 329 were in households with possible co-primary cases, 214 started follow-up >7 days after the index case’s illness onset, 137 had <2 follow-up specimens tested, 19 did not provide vaccination information, and 55 did not provide information on prior SARS-CoV-2 infection; **Supplemental table 2**), there were 1,532 household contacts from 905 households included in the analysis (**Figure 1**). Of these, 80% were enrolled from the sentinel approach and 63% were enrolled May–Nov 2022 when Omicron BA.4/5 predominated in the United States (**Supplemental figure 3)**. The median household size was 4 people (IQR: 2–4 people), 62% of household contacts were non-Hispanic White, 23% were Hispanic/Latino, 52% were female, and 28% were aged <18 years (**Table 1**).

**Figure 1:**
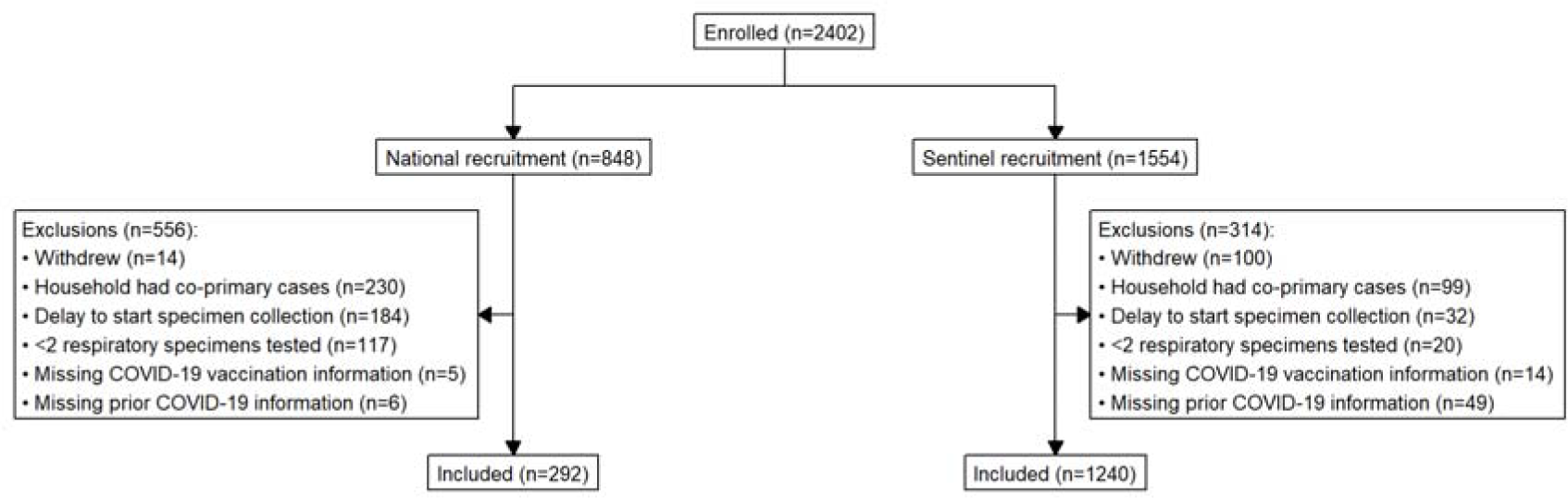
Eligibility and inclusion of household contacts in analysis of COVID-19 household transmission, United States, Sep 2021 – May 2023.

**Table 1:** Demographic characteristics of household contacts enrolled into case-ascertained household transmission studies of SARS-CoV-2 — United States, Sep 2021–May 2023.

|  | Overall (n=1532) | National approach (n=292) | Sentinel approach (n=1240) |
| --- | --- | --- | --- |
| **Time period, predominant SARS-CoV-2 variant (%)** |  |  |  |
| Sep-Nov 2021, Delta | 28 (1.8) | 0 (0.0) | 28 (2.3) |
| Dec 2021-Apr 2022, Omicron BA1/2 | 209 (13.6) | 17 (5.8) | 192 (15.5) |
| May-Nov 2022, Omicron BA4,5 | 962 (62.8) | 275 (94.2) | 687 (55.4) |
| Dec 2022-Apr 2023, XBB | 333 (21.7) | 0 (0.0) | 333 (26.9) |
| **Household size (%)** |  |  |  |
| <4 people | 772 (50.4) | 163 (55.8) | 609 (49.1) |
| 4+ people | 760 (49.6) | 129 (44.2) | 631 (50.9) |
| **Median people per bedroom [IQR]** | 1.25 [1.00, 1.67] | 1.00 [0.67, 1.25] | 1.25 [1.00, 1.67] |
| **Race-ethnicity (%)** |  |  |  |
| Asian, non-Hispanic | 69 (4.6) | 19 (6.5) | 50 (4.1) |
| Black, non-Hispanic | 85 (5.6) | 6 (2.1) | 79 (6.5) |
| Hispanic/Latino | 364 (24.0) | 29 (9.9) | 335 (27.4) |
| Multiple race, non-Hispanic | 43 (2.8) | 3 (1.0) | 40 (3.3) |
| Native Hawaiian or other Pacific Islander, non-Hispanic | 2 (0.1) | 0 (0.0) | 2 (0.2) |
| White, non-Hispanic | 934 (61.6) | 232 (79.5) | 702 (57.4) |
| Unknown/Refused | 19 (1.3) | 3 (1.0) | 16 (1.3) |
| **Self-reported sex (%)** |  |  |  |
| Female | 799 (52.2) | 139 (47.6) | 660 (53.2) |
| Male | 721 (47.1) | 144 (49.3) | 577 (46.5) |
| Non-binary/transgender | 12 (0.8) | 9 (3.1) | 3 (0.2) |
| **Age (years) (%)** |  |  |  |
| 0-4 | 92 (6.0) | 6 (2.1) | 86 (6.9) |
| 5-11 | 173 (11.3) | 37 (12.7) | 136 (11.0) |
| 12-17 | 161 (10.5) | 23 (7.9) | 138 (11.1) |
| 18-49 | 708 (46.2) | 126 (43.2) | 582 (46.9) |
| 50-64 | 254 (16.6) | 61 (20.9) | 193 (15.6) |
| 65+ | 144 (9.4) | 39 (13.4) | 105 (8.5) |
| **Prior COVID-19\* (%)** |  |  |  |
| No | 948 (61.9) | 184 (63.0) | 764 (61.6) |
| Yes | 584 (38.1) | 108 (37.0) | 476 (38.4) |
| **COVID-19 vaccination\*\* (%)** |  |  |  |
| No | 260 (17.0) | 18 (6.2) | 241 (19.4) |
| Yes | 1229 (80.2) | 272 (93.2) | 958 (77.3) |
| Ineligible | 43 (2.8) | 2 (0.7) | 41 (3.3) |
| **Immunity (%)** |  |  |  |
| No immunity | 167 (10.9) | 8 (2.7) | 159 (12.8) |
| Prior infection only | 130 (8.5) | 10 (3.4) | 119 (9.6) |
| Prior vaccination only | 778 (50.8) | 175 (59.9) | 603 (48.6) |
| Prior infection & vaccination | 451 (29.4) | 97 (33.2) | 355 (28.6) |
| No prior infection, vaccination unknown | 3 (0.2) | 1 (0.3) | 2 (0.2) |
| Prior infection, vaccination unknown | 3 (0.2) | 1 (0.3) | 2 (0.2) |

\* Prior COVID-19 was defined as anti-nucleocapsid antibodies detected at enrollment or plausibly self-reported prior positive test (with year of positive test reported).
\*\* COVID-19 vaccination was defined as $\geq 2$ verified or plausibly reported doses received more than 14 days prior to index case symptom onset.

### COVID-19 vaccination

Most household contacts (1,229/1,532, 80%) had received ≥2 doses of COVID-19 vaccine at least 14 days before their index case’s illness onset date, 3% (43/1,532) were age-ineligible to be vaccinated, and 17% (260/1,532) were age-eligible for vaccination but had received <2 doses (of which 79%, 206/260, had received 0 doses). Of those with ≥2 doses, 58% (708/1,272) had received ≥3 doses and only monovalent vaccine, 32% (402/1,272) had received 2 doses and only monovalent vaccine, 9% (119/1,272) had received ≥3 doses including ≥1 dose of bivalent vaccine, and 1 participant had received 2 doses including 1 bivalent dose. Among those who had received ≥2 doses, the median months since between last vaccination and index case illness onset was 208 (IQR: 114–309) days.

### Prior SARS-CoV-2 infection

Enrollment serology data were missing for 33% (503/1,532) of household contacts. Among the 1,011 household contacts with available antibody test results, 25% (262/1,029) had anti-nucleocapsid antibodies detected at enrollment. Five hundred eighty-four household contacts (38% of 1,532) reported a prior positive SARS-CoV-2 test or had anti-nucleocapsid antibodies detected at enrollment; of which, 22% (121/554) self-reported a prior positive test and had anti-nucleocapsid antibodies detected, 29% (158/554) self-reported a prior positive test but had negative results for anti-nucleocapsid antibodies 30% (167/554) self-reported a prior positive test and were not tested for antibodies, and 19% (82/554) had anti-nucleocapsid antibodies detected but did not report a prior positive test.

### Immunity

Of included household contacts, 11% (172/1,518) had <2 doses of COVID-19 vaccine and no evidence of prior infection, 8% (122/1,518) had prior SARS-CoV-2 infection but <2 doses of COVID-19 vaccine, 52% (792/1,518) had received ≥2 doses of COVID-19 vaccine but had no prior infection, and 28% (432/1,518) had received ≥2 doses of vaccine and had prior infection (**Table 2**). Those without immunity to COVID-19 (from either vaccination or prior infection) were more likely than those with some immunity to have been recruited in the sentinel approach, be children, and be from larger households (Table 2).

**Table 2:**
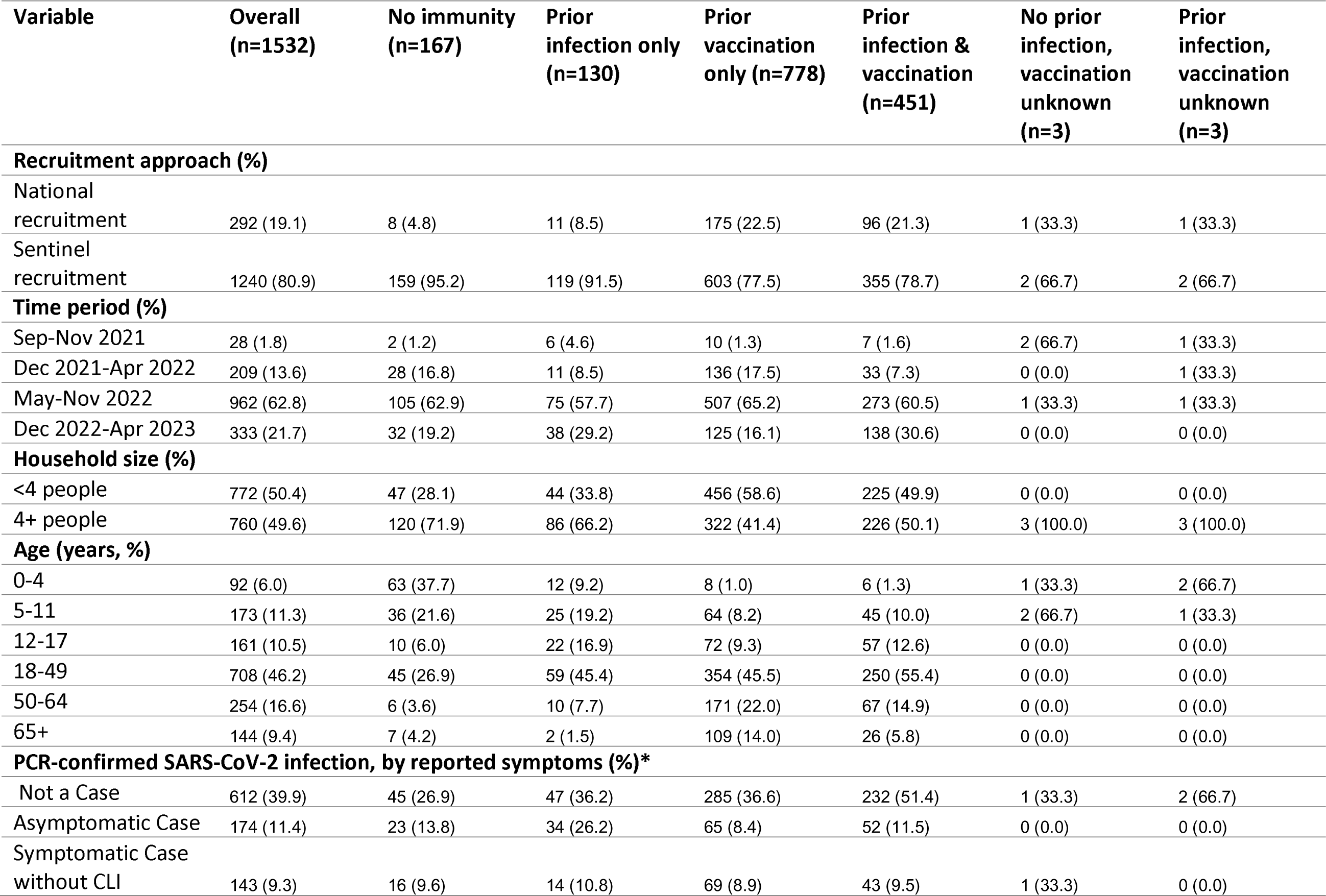
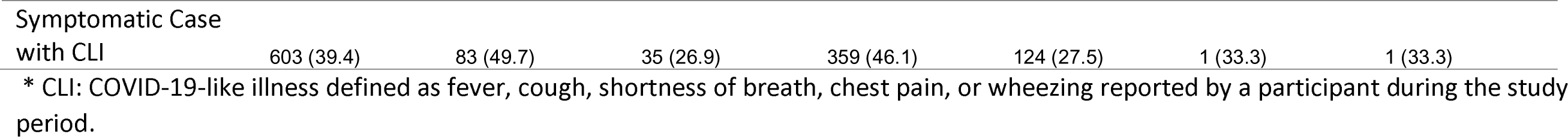
Demographic characteristics of included household contacts by COVID-19 vaccination and prior COVID-19 — case-ascertained household transmission study of SARS-CoV-2, United States, Sep 2021–May 2023.

| Variable | Overall<br>(n=1532) | No immunity<br>(n=167) | Prior<br>infection only<br>(n=130) | Prior<br>vaccination<br>only (n=778) | Prior<br>infection &<br>vaccination<br>(n=451) | No prior<br>infection,<br>vaccination<br>unknown<br>(n=3) | Prior<br>infection,<br>vaccination<br>unknown<br>(n=3) |
| --- | --- | --- | --- | --- | --- | --- | --- |
| **Recruitment approach (%)** |  |  |  |  |  |  |  |
| National<br>recruitment | 292 (19.1) | 8 (4.8) | 11 (8.5) | 175 (22.5) | 96 (21.3) | 1 (33.3) | 1 (33.3) |
| Sentinel<br>recruitment | 1240 (80.9) | 159 (95.2) | 119 (91.5) | 603 (77.5) | 355 (78.7) | 2 (66.7) | 2 (66.7) |
| **Time period (%)** |  |  |  |  |  |  |  |
| Sep-Nov 2021 | 28 (1.8) | 2 (1.2) | 6 (4.6) | 10 (1.3) | 7 (1.6) | 2 (66.7) | 1 (33.3) |
| Dec 2021-Apr 2022 | 209 (13.6) | 28 (16.8) | 11 (8.5) | 136 (17.5) | 33 (7.3) | 0 (0.0) | 1 (33.3) |
| May-Nov 2022 | 962 (62.8) | 105 (62.9) | 75 (57.7) | 507 (65.2) | 273 (60.5) | 1 (33.3) | 1 (33.3) |
| Dec 2022-Apr 2023 | 333 (21.7) | 32 (19.2) | 38 (29.2) | 125 (16.1) | 138 (30.6) | 0 (0.0) | 0 (0.0) |
| **Household size (%)** |  |  |  |  |  |  |  |
| <4 people | 772 (50.4) | 47 (28.1) | 44 (33.8) | 456 (58.6) | 225 (49.9) | 0 (0.0) | 0 (0.0) |
| 4+ people | 760 (49.6) | 120 (71.9) | 86 (66.2) | 322 (41.4) | 226 (50.1) | 3 (100.0) | 3 (100.0) |
| **Age (years, %)** |  |  |  |  |  |  |  |
| 0-4 | 92 (6.0) | 63 (37.7) | 12 (9.2) | 8 (1.0) | 6 (1.3) | 1 (33.3) | 2 (66.7) |
| 5-11 | 173 (11.3) | 36 (21.6) | 25 (19.2) | 64 (8.2) | 45 (10.0) | 2 (66.7) | 1 (33.3) |
| 12-17 | 161 (10.5) | 10 (6.0) | 22 (16.9) | 72 (9.3) | 57 (12.6) | 0 (0.0) | 0 (0.0) |
| 18-49 | 708 (46.2) | 45 (26.9) | 59 (45.4) | 354 (45.5) | 250 (55.4) | 0 (0.0) | 0 (0.0) |
| 50-64 | 254 (16.6) | 6 (3.6) | 10 (7.7) | 171 (22.0) | 67 (14.9) | 0 (0.0) | 0 (0.0) |
| 65+ | 144 (9.4) | 7 (4.2) | 2 (1.5) | 109 (14.0) | 26 (5.8) | 0 (0.0) | 0 (0.0) |
| **PCR-confirmed SARS-CoV-2 infection, by reported symptoms (%)\*** |  |  |  |  |  |  |  |
| Not a Case | 612 (39.9) | 45 (26.9) | 47 (36.2) | 285 (36.6) | 232 (51.4) | 1 (33.3) | 2 (66.7) |
| Asymptomatic Case | 174 (11.4) | 23 (13.8) | 34 (26.2) | 65 (8.4) | 52 (11.5) | 0 (0.0) | 0 (0.0) |
| Symptomatic Case<br>without CLI | 143 (9.3) | 16 (9.6) | 14 (10.8) | 69 (8.9) | 43 (9.5) | 1 (33.3) | 0 (0.0) |
| Symptomatic Case<br>with CLI | 603 (39.4) | 83 (49.7) | 35 (26.9) | 359 (46.1) | 124 (27.5) | 1 (33.3) | 1 (33.3) |

\* CLI: COVID-19-like illness defined as fever, cough, shortness of breath, chest pain, or wheezing reported by a participant during the study period.

### SARS-CoV-2 infections in household contacts

SARS-CoV-2 infections were detected in 60% (920/1,532) of contacts during follow-up. Of infected household contacts, 746 (81%) reported at least one symptom and 10% of symptomatic contacts (76/746) sought some form of medical care for their illness (including 3 who sought care at a hospital, 3 at an emergency department, 15 at an urgent care clinic, 24 at an outpatient clinic, 11 at a telehealth provider, and 13 who sought other care; care could have been sought at multiple locations). Infections were detected among 73% (122/167) of contacts with no immunity, 64% (83/130) of contacts with prior infection only, 63% (493/778) of contacts with prior vaccination only, and 49% (219/451) of contacts with hybrid immunity. Among those who had received at least 2 COVID-19 vaccine doses, there were fewer infections among household contacts with prior SARS-CoV-2 infection than those without (**Supplemental figure 4).** Compared with household contacts with no immunity, the risk of SARS-CoV-2 infection was not significantly reduced in contacts with vaccination only or those with prior SARS-CoV-2 infection only (**Table 3**). However, contacts with hybrid immunity (vaccination and prior SARS-CoV-2 infection) had significantly reduced risk of infection compared with contacts with no immunity (adjusted relative risk [aRR]: 0.81, 95% confidence interval [CI]: 0.70, 0.93, **Table 3**). In an analysis that examined the recency of both immunizing events together, the risk of infection was lower when the last immunizing event (vaccination and infection) occurred ≤6 months before index illness onset (aRR: 0.69, 95% CI: 0.57, 0.83) compared to no immunity. The adjusted risk of infection was lowest among those with hybrid immunity whose last immunizing event ≤6 months before the index illness onset, compared with all others (**Table 3**).

**Table 3:** Relative: Relative risk (RR) of SARS-CoV-2 infection among household contacts by COVID-19 vaccination or prior COVID-19.

|  | Contacts | Infections | Crude infection risk | Univariate RR | Adjusted Risk | Adjusted RR* |
| --- | --- | --- | --- | --- | --- | --- |
| **Immunity** |  |  |  |  |  |  |
| No immunity | 170 | 124 | 72.9% (65.8%, 79.1%) | Reference | 51.8% (44.6%, 60.2%) | Reference |
| Prior vaccination only | 778 | 493 | 63.4% (59.9%, 66.7%) | 0.90 (0.81, 0.99) | 55.2% (50.0%, 60.9%) | 1.02 (0.90, 1.15) |
| Prior infection only | 133 | 84 | 63.2% (54.7%, 70.9%) | 0.92 (0.81, 1.05) | 49.8% (42.1%, 58.9%) | 0.95 (0.84, 1.09) |
| Prior infection & vaccination | 451 | 219 | 48.6% (44.0%, 53.2%) | 0.73 (0.65, 0.82) | 41.8% (36.8%, 47.4%) | 0.81 (0.70, 0.93) |
| **Months since most recent immunizing event** |  |  |  |  |  |  |
| No immunity | 167 | 122 | 73.1% (65.8%, 79.2%) | Reference | 52.6% (45.3%, 60.9%) | Reference |
| Prior infection/vaccination:<br>last event >6 months ago | 456 | 293 | 64.3% (59.8%, 68.5%) | 0.90 (0.81, 1.00) | 55.7% (49.8%, 62.3%) | 1.02 (0.90, 1.15) |
| Prior infection/vaccination:<br>last event ≤6 months ago | 423 | 259 | 61.2% (56.5%, 65.8%) | 0.86 (0.77, 0.96) | 52.6% (47.0%, 58.8%) | 0.95 (0.84, 1.08) |
| Prior infection & vaccination:<br>last event >6 months ago | 230 | 128 | 55.7% (49.2%, 61.9%) | 0.81 (0.71, 0.92) | 48.7% (41.9%, 56.6%) | 0.91 (0.78, 1.06) |
| Prior infection & vaccination:<br>last event ≤6 months ago | 221 | 91 | 41.2% (34.9%, 47.8%) | 0.62 (0.52, 0.74) | 35.7% (29.9%, 42.6%) | 0.69 (0.57, 0.83) |
| Prior infection/vaccination:<br>last event time unknown | 29 | 24 | 82.8% (65.0%, 92.9%) | 1.15 (1.00, 1.31) | 61.5% (50.6%, 74.8%) | 1.11 (0.96, 1.29) |
| Unknown | 6 | 3 | 50.0% (18.8%, 81.2%) | 1.04 (0.85, 1.28) | 36.5% (17.3%, 76.8%) | 1.01 (0.79, 1.29) |

\* Model is adjusted for the recruitment approach for the household (national or sentinel), the age of the household contact (<5, 5–11, 12–17, 18–49, ≥50 years), household density (number of people/number of bedrooms), and month of enrollment (as a continuous variable from September 2021)

In secondary multivariable analyses that examined the separate role of vaccination history and prior infection on the risk of infection among household contacts, there was significantly reduced risk of infection among household contacts with prior SARS-CoV-2 infection (aRR: 0.84; 95% CI: 0.77-0.91) that was mainly driven by larger reductions in risk among those with prior SARS-CoV-2 infection in the previous 6 months (aRR: 0.70; 95% CI: 0.55-0.87; **Supplemental table 3**). In contrast, COVID-19 vaccination was not significantly associated with the risk of infection among household contacts (**Supplemental table 3**).

Results of planned sensitivity analyses showed that the estimated effects of vaccination alone and prior infection alone were consistent with those from the primary analyses and similar across different definitions of vaccination, prior infection, or when excluding households with episodes of COVID-19 in the 3 months prior to enrollment (**Supplemental tables 4-6)**. Results were also similar across different definitions of hybrid immunity and when participants with 1 or more doses of COVID-19 were considered vaccinated in the definition of hybrid immunity (**Supplemental tables 7-8**). Participants who had received a bivalent COVID-19 vaccine and had prior SARS-CoV-2 infection (hybrid immunity with a bivalent vaccine) had lower risk of infection compared with contacts with no immunity (aRR: 0.60, 95% CI: 0.40-0.90), which was similar to the relative risk among those with hybrid immunity who received their last immunizing event within 6 months of enrollment (**Supplemental table 9**). Results restricted to the sentinel approach were also consistent with the primary analysis (**Supplemental table 10**).

## Conclusions

We conducted two large, multi-state case-ascertained household transmission studies over 22 months in the United States, which encompassed circulation of SARS-CoV-2 Delta variant and Omicron subvariants BA.1, BA.2, BA.4, BA.5, BQ.1.1, and the emergence of XBB.1.5. Through prospective follow-up and frequent systematic testing for SARS-CoV-2, household contacts with evidence of prior SARS-CoV-2 infection and COVID-19 vaccination had a significantly lower risk of infection compared to those without prior immunizing events. The protection conferred by hybrid immunity was highest when the last immunizing event (either vaccination or infection) had occurred in the previous 6 months, reducing the risk by 30%.

While population immunity against SARS-CoV-2 is evolving, it currently comprises a mixture of vaccine- and infection-induced immunity).^6, 20^ There is consistent evidence that vaccine effectiveness against severe disease and medically-attended SARS-CoV-2 infection is modulated by prior infection.^2, 10–12^ The present study found that the greatest protection against mild, largely non-medically-attended SARS-CoV-2 infection was afforded by hybrid immunity. The effect of hybrid immunity against infection, regardless of symptoms, acquired in household settings in the present study was modest, with a 14% reduction in risk of infection with any hybrid immunity and 30% reduction among those with an immunizing event in the past 6 months. Notably, the protective effect of hybrid immunity observed in this household study was similar to the protective effect observed in the Omicron era among incarcerated individuals who reside together in congregate settings^16^. This appears lower than effects against infection (61% among those with an immunizing event in the past 6 months) or severe disease (80% among those with an immunizing event in the past 6 months) in prior meta-regressions of other studies, which primarily examined symptomatic infections and were not conducted in close-contact settings.^9^ While, preventing severe, critical, and fatal outcomes is a main goal of COVID-19 prevention and vaccination programs, it is also important to understand the impact of vaccination and prior infection on milder illness and infection as most SARS-CoV-2 infections confer a cost from missed days of work or school and can result in sustained transmission.

Our study also highlights how transmissible SARS-CoV-2 can be in high-contact settings. Among household contacts with hybrid immunity who had their last immunizing event in the 6 months prior to their household being affected by SARS-CoV-2, 42% were infected with SARS-CoV-2 during follow-up. Nearly two-thirds of household contacts without immunity were infected. Additional prevention measures that can be used in these closed, high-interaction settings include wearing a high-quality mask and isolating promptly from others in the household if you suspect that you have COVID-19.^21^

This household transmission study is subject to multiple limitations. First, the selection criteria for analysis excluded a large proportion of households. While exclusions were based on lack of necessary data for determination of transmission events, excluded households differed in important ways, such as race-ethnicity, household size, and time period, which may limit the generalizability of study findings. Second, prior infection is difficult to ascertain accurately, as self-report and solely serologic definitions may both be affected by misclassification.^22^ However, our planned sensitivity analyses demonstrated that results based solely on self-report are comparable to those based solely on serology, suggesting that this potential misclassification did not introduce substantial bias. Third, vaccination may have been misclassified, by either self-report or by limited ability to accurately verify doses received. Nevertheless, sensitivity analyses around the definition of vaccination demonstrated similar results. Fourth, while this study was large for a household transmission study, some subgroups were small, which limited precision. Moreover, the number of infections among household contacts was insufficient to analyze the risk of severe disease, which was not common in this cohort. Finally, multiple variants of SARS-CoV-2 emerged over the course of the study, which limited our ability to estimate the effectiveness of vaccination against infections with specific variants.

In conclusion, this study, which included serological testing to confirm prior infection and daily PCR testing of all contacts regardless of symptoms, suggests the risk of mild SARS-CoV-2 infection is not eliminated but is significantly lower in household contacts who had prior infection and vaccination, compared with immunity from prior infection alone, vaccination alone, and no immunity. These findings underscore the importance of staying updated with COVID-19 vaccinations, even for individuals with prior infection.

## Data Availability

All data produced in the present study are available upon reasonable request to the authors

## In addition, we would like to acknowledge

### Marshfield Clinic Research Institute

Hannah Berger, Brianna Breu, Gina Burbey, Leila Deering, DeeAnn Hertel, Garrett Heuer, Sarah Kopitzke, Carrie Marcis, Jennifer Meece, Vicki Moon, Jennifer Moran, Miriah Rotor, Carla Rottscheit, Elisha Stefanski, Sandy Strey, and Melissa Strupp.

### Vanderbilt University Medical Center

Judy King, Lauren Milner, Andrea Stafford Hintz, Samuel Massion, John Meghreblian, Brittany Creasman, Catalina Padilla-Azain, Daniel Chandler, Paige Yates, Jorge Celedonio, Ryan Dalforno, Brianna Schibley-Laird, Ruby Swaim, Erica Anderson, Suryakala Sarilla, Timothy Williams, Marcia Blair, Laura Short, Bryan Peterson, Christopher Lindsell, Zhouwen Liu, His-nien Tan, Rendie E. McHenry, Bryan P. M. Peterson.

### Disclaimer

The findings and conclusions in this report are those of the authors and do not necessarily represent the official position of the Centers for Disease Control and Prevention.

### Funding/Support

These research studies were supported the Centers for Disease Control and Prevention (CDC) by contracts 75D30121C11656 (jointly funded by CDC and the US Food and Drug Administration, awarded to Vanderbilt University Medical Center) and 75D30121C11571 (awarded to Westat).

### Access to Data

Dr. Rolfes and Dr. Mellis had full access to all of the data in the study and take responsibility for the integrity of the data and the accuracy of the data analysis.

### Additional Contributions

We acknowledge all of the households who agreed to participate in the study and the research coordinators at each of the participating enrollment sites.

---

**Supplemental Figure 1.**
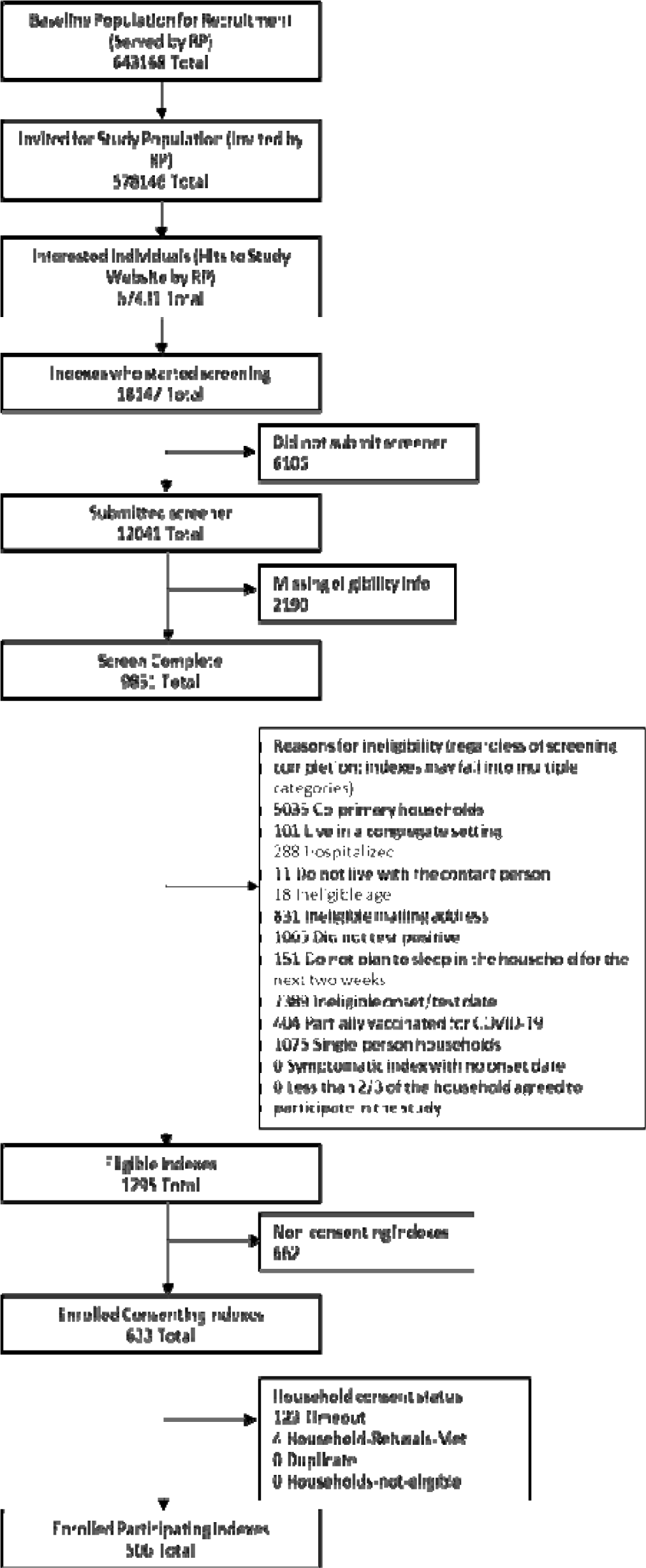
Reasons for ineligibility prior to enrollment, RVTN-National approach.

**Supplemental Figure 2:**
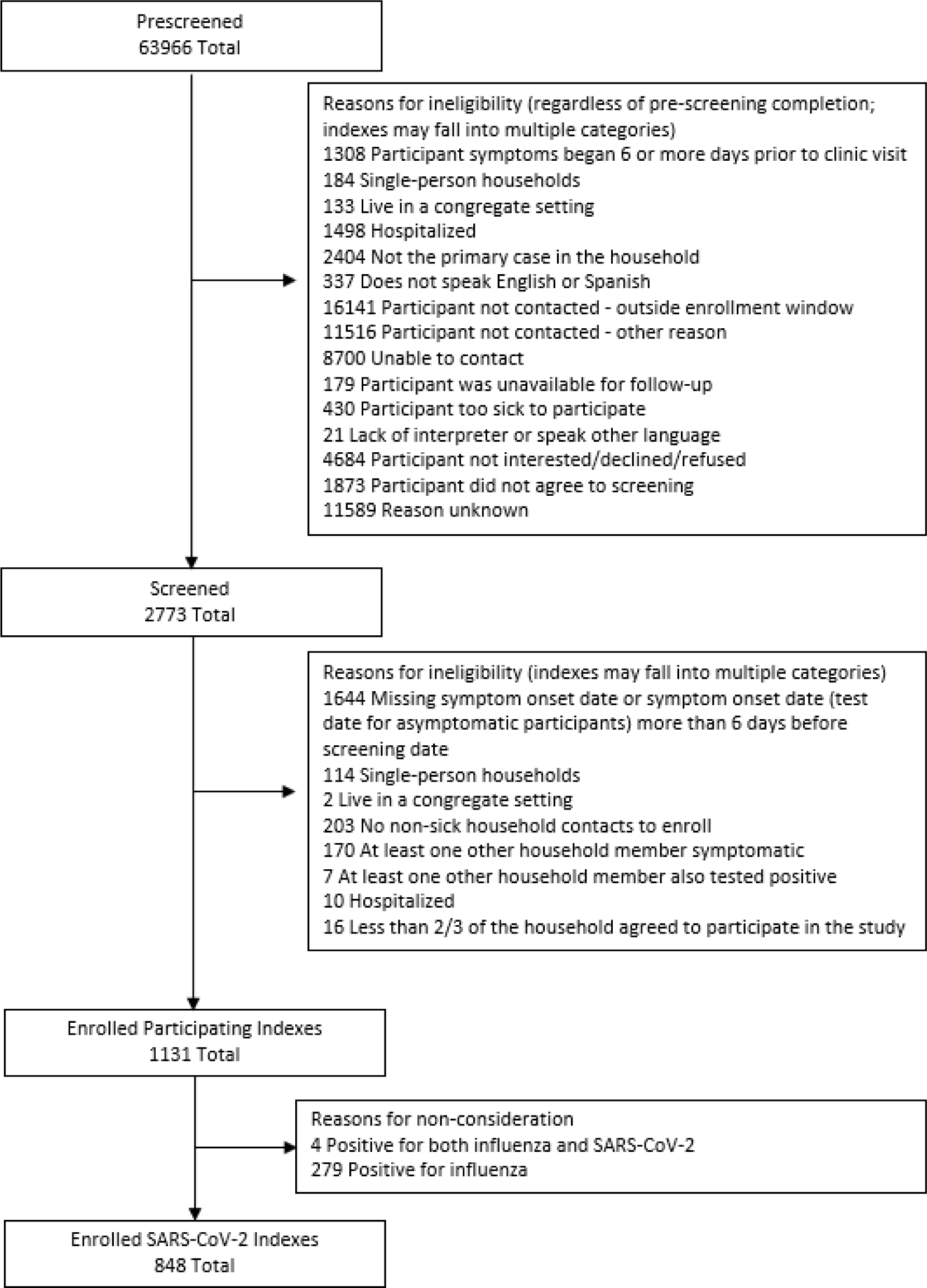
Reasons for ineligibility prior to enrollment, RVTN-Sentinel approach.

**Supplemental Figure 3:**
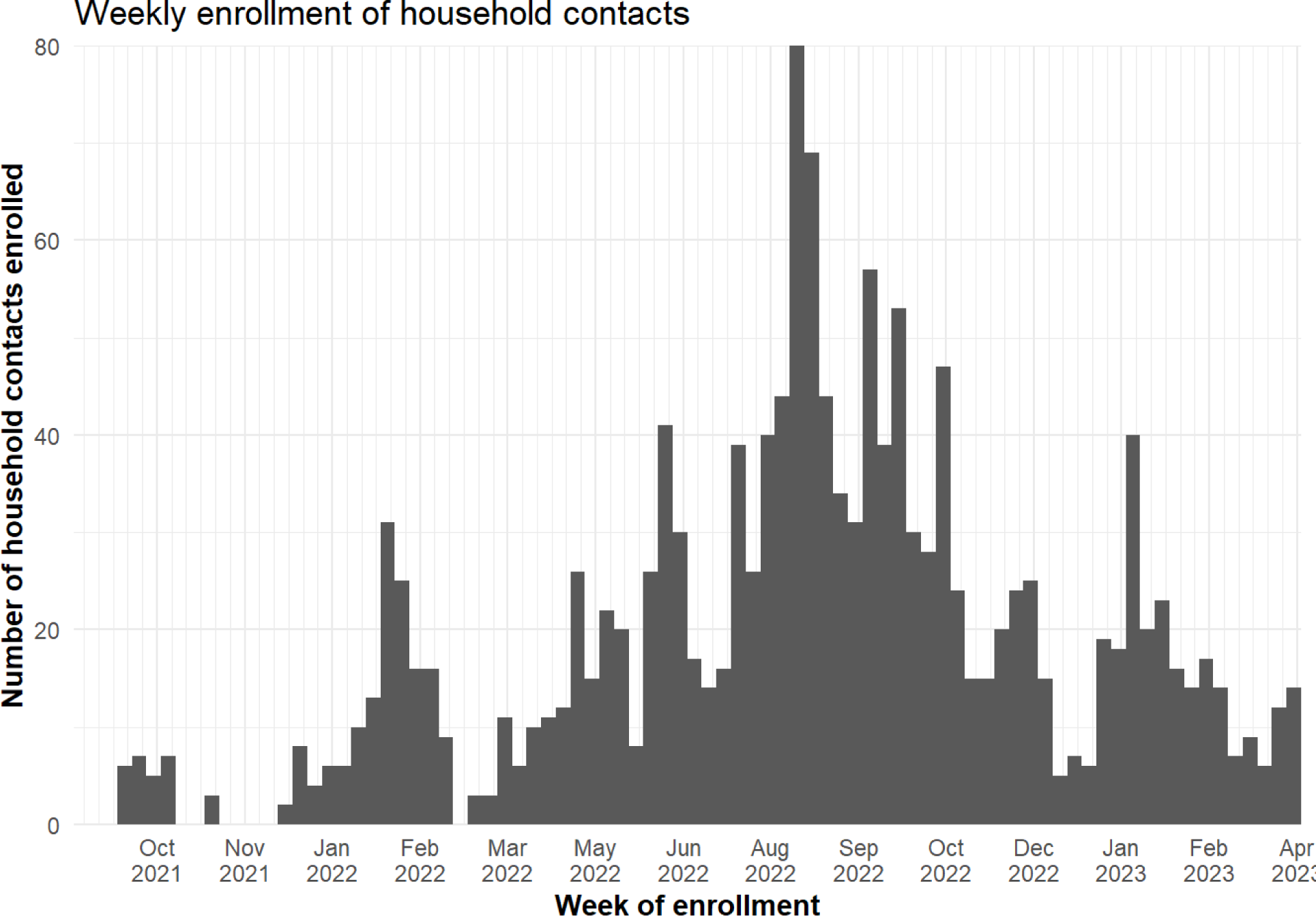
Number of households enrolled by week, September 2021—May 2023.

**Supplemental Figure 4:**
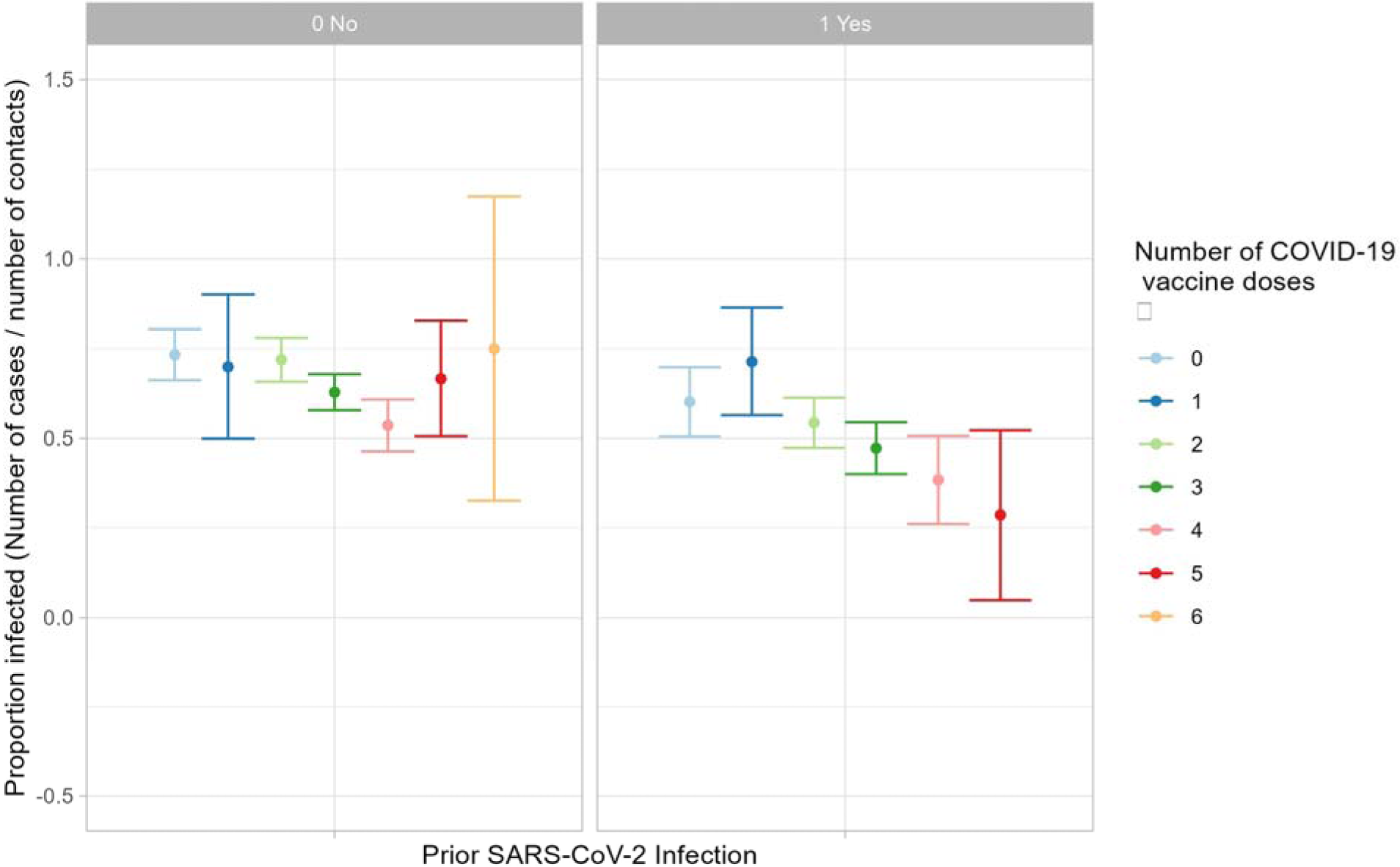
Unadjusted risk of SARS-CoV-2 infection among household contacts stratified by COVID-19 vaccination doses and prior SARS-CoV-2 infection, as defined for the initial analysis.

**Supplemental Table 1:** Sensitivity analyses considered.

| Exposure | Definition used in initial analysis | Definition used in sensitivity analysis | Supplemental Table |
| --- | --- | --- | --- |
| COVID-19 vaccination | ≥2 verified or plausibly reported doses | ≥2 verified doses | 4 |
| Prior SARS-CoV-2 infection | Anti-nucleocapsid antibodies detected at enrollment or plausibly self-reported prior positive test (with year of positive test reported) | Anti-nucleocapsid antibodies detected at enrollment; restricted to household contacts with complete antibody testing | 5 |
|  |  | Anti-nucleocapsid antibodies or self-reported prior positive test with month and year of test reported | 5 |
|  |  | Same as initial analysis but analysis is restricted analysis to households who did not report positive tests in any household members in the 3 months before enrollment | 6 |
| Immunity | Hybrid immunity: anti-nucleocapsid antibodies or plausibly reported prior positive test and ≥2 verified or plausibly reported COVID-19 vaccine doses<br>Only vaccination: ≥2 verified vaccination doses and no anti-nucleocapsid antibodies and no plausibly reported prior positive test<br>Only prior SARS-CoV-2 infection: anti-nucleocapsid antibodies or plausibly reported prior positive test and <2 verified or plausibly reported COVID-19 vaccine doses<br>No immunity: <2 verified or plausibly reported COVID-19 vaccine doses and no detected anti-nucleocapsid antibodies and no plausibly reported prior positive test | Hybrid immunity: anti-nucleocapsid antibodies and ≥2 verified COVID-19 vaccine doses,<br>Only vaccination: ≥2 verified vaccination doses and no detected anti-nucleocapsid antibodies<br>Only prior SARS-CoV-2 infection: detected anti-nucleocapsid antibodies and <2 verified doses of COVID-19 vaccination<br>No immunity: <2 verified doses of COVID-19 vaccine and no detected anti-nucleocapsid antibodies | 7 |
|  |  | Hybrid immunity: anti-nucleocapsid antibodies or plausibly reported prior<br>positive test and $\geq 1$ verified or plausibly reported COVID-19 vaccine doses<br>Only vaccination: $\geq 1$ verified vaccination doses and no anti-nucleocapsid antibodies and no plausibly reported prior positive test<br>Only prior SARS-CoV-2 infection: anti-nucleocapsid antibodies or plausibly reported prior positive test and 0 verified or plausibly reported COVID-19 vaccine doses<br>No immunity: 0 verified or plausibly reported COVID-19 vaccine doses and no detected anti-nucleocapsid antibodies and no plausibly reported prior positive test | 8 |
|  |  | Hybrid immunity, bivalent dose: anti-nucleocapsid antibodies or plausibly reported prior positive test and $\geq 2$ verified or plausibly reported COVID-19 vaccine doses (including 1 or more bivalent dose)<br>Hybrid immunity, monovalent only: anti-nucleocapsid antibodies or plausibly reported prior positive test and $\geq 2$ verified or plausibly reported COVID-19 vaccine doses (0 bivalent doses)<br>Only vaccination, bivalent dose: $\geq 2$ verified vaccination doses (1 or more bivalent doses) and no anti-nucleocapsid antibodies and no plausibly reported prior positive test<br>Only monovalent vaccination: $\geq 2$ verified vaccination doses (0 bivalent doses) and no anti-nucleocapsid antibodies and no plausibly reported prior positive test<br>Only prior SARS-CoV-2 infection: anti-nucleocapsid antibodies or plausibly<br>reported prior positive test and <2 verified or plausibly reported COVID-19 vaccine doses<br>No immunity: <2 verified or plausibly reported COVID-19 vaccine doses and no detected anti-nucleocapsid antibodies and no plausibly reported prior positive test | 9 |

**Supplemental table 2:** Demographics of enrolled but analytically excluded household contacts, compared to analytically included household contacts.

|  | Overall (n=2402) | Included (n=1532) | Excluded (n=870) | P |
| --- | --- | --- | --- | --- |
| **Median household size [IQR]** | 4.00 [2.00, 4.00] | 3.00 [2.00, 4.00] | 4.00 [3.00, 4.00] | 0.002 |
| **Median people per bedroom [IQR]** | 1.20 [1.00, 1.50] | 1.25 [1.00, 1.67] | 1.00 [1.00, 1.50] | 0.215 |
| **Time period (%)** |  |  |  |  |
| Sep-Nov 2021 | 35 ( 1.5) | 28 ( 1.8) | 7 ( 0.8) | <0.001 |
| Dec 2021-Apr 2022 | 298 (12.4) | 209 (13.6) | 89 (10.3) |  |
| May-Nov 2022 | 1669 (69.7) | 962 (62.8) | 707 (81.9) |  |
| Dec 2022-Apr 2023 | 393 (16.4) | 333 (21.7) | 60 ( 7.0) |  |
| **Race-ethnicity (%)** |  |  |  |  |
| Asian, Non-Hispanic | 118 ( 5.1) | 69 ( 4.6) | 49 ( 6.3) | 0.339 |
| Black, Non-Hispanic | 119 ( 5.2) | 85 ( 5.6) | 34 ( 4.4) |  |
| Hispanic/Latino | 551 (24.0) | 364 (24.0) | 187 (24.0) |  |
| Multiple race, Non-Hispanic | 63 ( 2.7) | 43 ( 2.8) | 20 ( 2.6) |  |
| NH/OPI, Non-Hispanic | 4 ( 0.2) | 2 ( 0.1) | 2 ( 0.3) |  |
| Unknown/Refused | 34 ( 1.5) | 19 ( 1.3) | 15 ( 1.9) |  |
| White, Non-Hispanic | 1406 (61.3) | 934 (61.6) | 472 (60.6) |  |
| **Household size (%)** |  |  |  |  |
| <4 people | 1182 (49.2) | 772 (50.4) | 410 (47.2) | 0.142 |
| 4+ people | 1219 (50.8) | 760 (49.6) | 459 (52.8) |  |
| **Age (years) %** |  |  |  |  |
| 0-4 | 157 ( 6.5) | 92 ( 6.0) | 65 ( 7.5) | 0.182 |
| 5-11 | 297 (12.4) | 173 (11.3) | 124 (14.3) |  |
| 12-17 | 254 (10.6) | 161 (10.5) | 93 (10.7) |  |
| 18-49 | 1082 (45.0) | 708 (46.2) | 374 (43.0) |  |
| 50-64 | 388 (16.2) | 254 (16.6) | 134 (15.4) |  |
| 65+ | 224 ( 9.3) | 144 ( 9.4) | 80 ( 9.2) |  |
| **Self-reported sex (%)** |  |  |  |  |
| Female | 1212 (51.0) | 799 (52.2) | 413 (48.8) | 0.286 |
| Male | 1146 (48.2) | 721 (47.1) | 425 (50.2) |  |
| Non-binary/transgender | 20 ( 0.8) | 12 ( 0.8) | 8 ( 0.9) |  |
| **Prior COVID-19 infection (%)** |  |  |  |  |
| No | 1388 (57.8) | 948 (61.9) | 440 (50.6) | <0.001 |
| Yes | 863 (35.9) | 584 (38.1) | 279 (32.1) |  |
| Unknown | 151 ( 6.3) | 0 ( 0.0) | 151 (17.4) |  |
| **Baseline serostatus (%)** |  |  |  |  |
| Anti-N negative | 986 (41.0) | 767 (50.1) | 219 (25.2) | <0.001 |
| Anti-N positive | 381 (15.9) | 262 (17.1) | 119 (13.7) |  |
| Unknown | 1035 (43.1) | 503 (32.8) | 532 (61.1) |  |
| **Plausibly self-reported prior infection (%)** |  |  |  |  |
| No, prior COVID | 1388 (57.8) | 948 (61.9) | 440 (50.6) | <0.001 |
| Yes, prior COVID | 863 (35.9) | 584 (38.1) | 279 (32.1) |  |
| Unknown, prior COVID | 151 ( 6.3) | 0 ( 0.0) | 151 (17.4) |  |
| **SARS-CoV-2 vaccination status (%)** |  |  |  |  |
| Unvaccinated, <2 doses | 407 (16.9) | 260 (17.0) | 147 (16.9) | <0.001 |
| Vaccinated, last dose <120 days | 481 (20.0) | 334 (21.8) | 147 (16.9) |  |
| Vaccinated, last dose 121-180 days | 284 (11.8) | 207 (13.5) | 77 ( 8.9) |  |
| Vaccinated, last dose >180 days | 1079 (44.9) | 688 (44.9) | 391 (44.9) |  |
| Ineligible | 70 ( 2.9) | 43 ( 2.8) | 27 ( 3.1) |  |
| Unknown | 81 ( 3.4) | 0 ( 0.0) | 81 ( 9.3) |  |
| **Hybrid immune status (%)** |  |  |  |  |
| No immunity | 407 (16.9) | 260 (17.0) | 147 (16.9) | <0.001 |
| Prior infection only | 481 (20.0) | 334 (21.8) | 147 (16.9) |  |
| Prior vaccination only | 284 (11.8) | 207 (13.5) | 77 ( 8.9) |  |
| Hybrid immunity | 1079 (44.9) | 688 (44.9) | 391 (44.9) |  |
| No prior infection, Vax Unk | 70 ( 2.9) | 43 ( 2.8) | 27 ( 3.1) |  |
| Prior infection, Vax Unk | 81 ( 3.4) | 0 ( 0.0) | 81 ( 9.3) |  |
| Prior infection Unk, No vaccine | 37 ( 1.5) | 0 ( 0.0) | 37 ( 4.3) |  |
| Prior infection Unk, Vaccination | 71 ( 3.0) | 0 ( 0.0) | 71 ( 8.2) |  |
| Prior infection Unk, Vax Unk | 43 ( 1.8) | 0 ( 0.0) | 43 ( 4.9) |  |
| **Symptom status (%)** |  |  |  |  |
| No symptoms | 719 (29.9) | 436 (28.5) | 283 (32.5) | 0.041 |
| Symptoms | 1683 (70.1) | 1096 (71.5) | 587 (67.5) |  |
| Unk. | 719 (29.9) | 436 (28.5) | 283 (32.5) | 0.041 |

**Supplemental table 3:** Relative risk (RR) of infection among household contacts by prior SARS-CoV-2 infection or COVID-19 vaccination, not accounting for hybrid immune status.

|  | Contacts | Infections | Crude infection risk | Univariate RR | Adjusted RR |
| --- | --- | --- | --- | --- | --- |
| **COVID-19 Vaccination** |  |  |  |  |  |
| No | 260 | 175 | 67.3% (61.4%, 72.7%) | Reference | Reference |
| Yes | 1229 | 712 | 57.9% (55.2%, 60.7%) | 0.87 (0.79, 0.97) | 0.95 (0.85, 1.06) |
| Ineligible | 43 | 33 | 76.7% (62.1%, 87.0%) | 1.09 (0.93, 1.28) | 1.07 (0.90, 1.26) |
| **Days since last COVID-19 vaccine dose** |  |  |  |  |  |
| Unvaccinated, <2 doses | 260 | 175 | 67.3% (61.4%, 72.7%) | Reference | Reference |
| Vaccinated, last dose >180 days | 688 | 415 | 60.3% (56.6%, 63.9%) | 0.90 (0.81, 1.00) | 1.00 (0.89, 1.13) |
| Vaccinated, last dose <120 days | 334 | 185 | 55.4% (50.0%, 60.6%) | 0.83 (0.73, 0.95) | 0.89 (0.77, 1.01) |
| Vaccinated, last dose 121-180 days | 207 | 112 | 54.1% (47.3%, 60.8%) | 0.81 (0.69, 0.94) | 0.87 (0.75, 1.01) |
| Ineligible | 43 | 33 | 76.7% (62.1%, 87.0%) | 1.09 (0.93, 1.27) | 1.04 (0.89, 1.23) |
| **Prior COVID-19** |  |  |  |  |  |
| No, prior COVID | 948 | 617 | 65.1% (62.0%, 68.1%) | Reference | Reference |
| Yes, prior COVID | 584 | 303 | 51.9% (47.8%, 55.9%) | 0.84 (0.77, 0.91) | 0.83 (0.77, 0.90) |
| **Months since prior COVID-19** |  |  |  |  |  |
| No prior COVID | 989 | 631 | 63.8% (60.8%, 66.7%) | Reference | Reference |
| Prior COVID, unknown time | 317 | 163 | 51.4% (45.9%, 56.9%) | 0.83 (0.74, 0.93) | 0.83 (0.74, 0.93) |
| Prior COVID ≤6 months ago | 64 | 26 | 40.6% (29.4%, 52.9%) | 0.71 (0.56, 0.88) | 0.69 (0.55, 0.86) |
| Prior COVID >6 months ago | 46 | 26 | 56.5% (42.2%, 69.8%) | 0.96 (0.75, 1.23) | 0.98 (0.77, 1.24) |

**Supplemental table 4:** Sensitivity analyses of the impact of vaccination on infection risk by the definition of vaccination.

|  |  |
| --- | --- |
|  | Relative risk (vs. unvaccinated) |
| **≥2 doses of SARS-CoV-2 vaccine (primary analysis)** |  |
| Yes | 0.95 (0.85, 1.06) |
| Ineligible | 1.07 (0.90, 1.26) |
| **Sensitivity 1. Verified vaccines only (≥2 verified doses)** |  |
| Yes | 0.96 (0.87, 1.07) |
| Ineligible | 1.08 (0.91, 1.27) |
| Unknown | 1.27 (0.84, 1.91) |
| **Sensitivity 2. Considering ≥1 dose as vaccinated** |  |
| Yes | 0.98 (0.87, 1.10) |
| Ineligible | 1.08 (0.91, 1.27) |

**Supplemental Table 5:** Sensitivity analyses of the impact of prior infection on infection risk by the definition of prior infection.

| Level | Relative risk (vs. no prior infection) |
| --- | --- |
| **SARS-CoV-2 infection (primary analysis)** |  |
| Yes, prior COVID | 0.83 (0.77, 0.90) |
| **Sensitivity 1. Anti-N antibodies only** |  |
| Anti-Nucleocapsid antibodies | 0.87 (0.77, 0.98) |
| **Sensitivity 2. Plausible self-report also requires month** |  |
| Yes | 0.83 (0.77, 0.90) |

**Supplemental Table 6:** Sensitivity analyses of the impact of prior infection on infection risk, excluding households with other episodes of COVID-19 prior to enrollment.

| Level | Relative risk (vs. no infection) |
| --- | --- |
| **SARS-CoV-2 infection (primary analysis)** |  |
| Yes, prior COVID | 0.83 (0.77, 0.90) |
| **Sensitivity 1. Excluding households with other episodes within the past 3 months** |  |
| Yes, prior COVID | 0.85 (0.78, 0.92) |

**Supplemental Table 7:** Sensitivity analyses of the impact of hybrid immunity on infection risk by the definitions of vaccination and prior infection.

|  | Relative risk (vs no immunity) |
| --- | --- |
| **immunity (primary analysis)** |  |
| Prior infection only | 0.97 (0.84, 1.10) |
| Prior vaccination only | 1.04 (0.91, 1.18) |
| Prior infection & vaccination | 0.84 (0.72, 0.97) |
| No prior infection, vaccination unknown | 1.01 (0.70, 1.46) |
| Prior infection, vaccination unknown | 0.90 (0.57, 1.42) |
| **Sensitivity 1. Immunity (strict definitions)** |  |
| Prior infection only | 1.01 (0.86, 1.19) |
| Prior vaccination only | 0.97 (0.86, 1.09) |
| Hybrid immunity | 0.78 (0.64, 0.94) |
| No prior infection, Vaccination unknown | 0.93 (0.64, 1.36) |
| Prior infection, Vaccination unknown | 1.13 (0.59, 2.14) |
| Prior infection unknown, No vaccine | 0.95 (0.82, 1.10) |
| Prior infection unknown, Vaccination | 0.96 (0.83, 1.11) |
| Prior infection unknown, Vaccination unknown | 1.30 (0.90, 1.88) |
| **Time since last immunizing event (primary analysis)** |  |
| Prior infection or vaccination: last event ≤6 months ago | 0.96 (0.84, 1.09) |
| Prior infection or vaccination: last event >6 months ago | 1.04 (0.92, 1.18) |
| Prior infection or vaccination: last event time unknown | 1.13 (0.95, 1.34) |
| Prior infection & vaccination: last event ≤6 months ago | 0.72 (0.60, 0.87) |
| Prior infection & vaccination: last event >6 months ago | 0.93 (0.79, 1.09) |
| Unknown | 0.94 (0.70, 1.26) |
| **Sensitivity 1. Time since last immunizing event (strict definitions)** |  |
| Prior infection or vaccination: last event ≤6 months ago | 0.89 (0.78, 1.02) |
| Prior infection or vaccination: last event >6 months ago | 1.02 (0.90, 1.15) |
| Prior infection or vaccination: last event time unknown | 1.12 (0.92, 1.35) |
| Prior infection & vaccination: last event ≤6 months ago | 0.56 (0.39, 0.81) |
| Prior infection & vaccination: last event >6 months ago | 0.95 (0.79, 1.14) |
| Unknown | 0.96 (0.84, 1.09) |

**Supplemental Table 8:** Sensitivity analyses of the impact of hybrid immunity on infection risk, considering 1 or more doses of COVID-19 vaccine as vaccinated.

|  | Relative risk vs no immunity |
| --- | --- |
| **immunity (main analysis; vs. no immunity)** |  |
| Prior infection only | 0.97 (0.84, 1.10) |
| Prior vaccination only | 1.04 (0.91, 1.18) |
| Prior infection & vaccination | 0.84 (0.72, 0.97) |
| No prior infection, vaccination unknown | 1.01 (0.70, 1.46) |
| Prior infection, vaccination unknown | 0.90 (0.57, 1.42) |
| **Sensitivity 1. immunity (1 dose as vaccinated; vs. no immunity)** |  |
| Prior infection only | 0.95 (0.82, 1.11) |
| Prior vaccination only | 1.06 (0.94, 1.19) |
| Hybrid immunity | 0.86 (0.75, 0.99) |
| No prior infection, Vaccination unknown | 1.39 (1.17, 1.66) |
| Prior infection, Vaccination unknown | 0.83 (0.50, 1.37) |
| **Sensitivity 2. Months since last immunizing event (1 dose as vaccinated; vs. no immunity)** |  |
| Prior infection/vaccination: last event ≤6 months ago | 0.98 (0.87, 1.11) |
| Prior infection/vaccination: last event >6 months ago | 1.04 (0.93, 1.17) |
| Prior infection/vaccination: last event time unknown | 1.14 (0.95, 1.35) |
| Prior infection & vaccination: last event ≤6 months ago | 0.74 (0.62, 0.88) |
| Prior infection & vaccination: last event >6 months ago | 0.95 (0.82, 1.09) |
| Unknown | 1.02 (0.81, 1.30) |

**Supplemental Table 9:** Sensitivity analyses of the impact of hybrid immunity on infection risk, considering bivalent doses.

|  | Relative risk (vs no immunity) |
| --- | --- |
| **Immunity (primary analysis)** |  |
| Prior infection only | 0.97 (0.84, 1.10) |
| Prior vaccination only | 1.04 (0.91, 1.18) |
| Prior infection & vaccination | 0.84 (0.72, 0.97) |
| No prior infection, vaccination unknown | 1.01 (0.70, 1.46) |
| Prior infection, vaccination unknown | 0.90 (0.57, 1.42) |
| **Sensitivity 1. immunity, accounting for vaccine valence** |  |
| Prior infection only | 0.95 (0.83, 1.09) |
| Prior monovalent vaccination only | 1.03 (0.91, 1.16) |
| Prior bivalent vaccination only | 1.03 (0.83, 1.27) |
| Hybrid immunity, with monovalent vaccination | 0.85 (0.73, 0.98) |
| Hybrid immunity, with bivalent vaccination | 0.60 (0.40, 0.90) |
| No prior infection, Vax Unk | 1.38 (1.15, 1.67) |
| Prior infection, Vax Unk | 0.82 (0.49, 1.39) |

**Supplemental Table 10:** Sensitivity analyses of the relative risk (95% confidence interval) of infection, stratified by study approach.

|  | Combined | Sentinel approach | National approach |
| --- | --- | --- | --- |
| **SARS-CoV-2 vaccination (vs. 0-1 dose)** |  |  |  |
| Yes | 0.95 (0.85, 1.06) | 0.99 (0.89, 1.10) | 0.92 (0.83, 1.03) |
| Ineligible | 1.07 (0.90, 1.26) | 1.07 (0.91, 1.26) | 1.07 (0.91, 1.27) |
| **Time since last SARS-CoV-2 vaccination (vs. 0-1 dose)** |  |  |  |
| Vaccinated, last dose <120 days | 0.89 (0.77, 1.01) | 0.92 (0.81, 1.05) | 0.86 (0.75, 0.99) |
| Vaccinated, last dose 121-180 days | 0.87 (0.75, 1.01) | 0.93 (0.80, 1.08) | 0.85 (0.73, 0.99) |
| Vaccinated, last dose >180 days | 1.00 (0.89, 1.13) | 1.04 (0.93, 1.16) | 0.97 (0.87, 1.09) |
| Ineligible | 1.04 (0.89, 1.23) | 1.05 (0.90, 1.24) | 1.05 (0.89, 1.25) |
| **Plausibly self-reported prior infection (vs. no)** |  |  |  |
| Yes, prior COVID | 0.83 (0.77, 0.90) | 0.85 (0.78, 0.92) | 0.84 (0.78, 0.91) |
| **Time since plausibly self-reported prior infection (vs. no prior infection)** |  |  |  |
| Prior COVID <=6 months ago | 0.69 (0.55, 0.86) | 0.76 (0.62, 0.93) | 0.71 (0.57, 0.88) |
| Prior COVID >6 months ago | 0.98 (0.77, 1.24) | 0.95 (0.74, 1.23) | 1.00 (0.79, 1.26) |
| Prior COVID, unknown time | 0.83 (0.74, 0.93) | 0.81 (0.73, 0.91) | 0.85 (0.76, 0.94) |
| **Hybrid immunity (vs. no immunity)** |  |  |  |
| Prior infection only | 0.95 (0.84, 1.09) | 0.96 (0.85, 1.09) | 0.94 (0.83, 1.06) |
| Prior vaccination only | 1.02 (0.90, 1.15) | 1.05 (0.93, 1.19) | 0.95 (0.86, 1.06) |
| Prior infection & vaccination | 0.81 (0.70, 0.93) | 0.85 (0.74, 0.98) | 0.76 (0.67, 0.86) |
| **Time since last immunizing event (vs. no immunity)** |  |  |  |
| Prior infection/vaccination: last event <=6 months ago | 0.95 (0.84, 1.08) | 1.00 (0.88, 1.13) | 0.92 (0.81, 1.05) |
| Prior infection/vaccination: last event >6 months ago | 1.02 (0.90, 1.15) | 1.01 (0.90, 1.15) | 0.99 (0.87, 1.12) |
| Prior infection/vaccination: last event time unknown | 1.11 (0.96, 1.29) | 1.14 (0.99, 1.31) | 1.13 (0.98, 1.31) |
| Prior infection & vaccination: last event <=6 months ago | 0.69 (0.57, 0.83) | 0.72 (0.60, 0.87) | 0.68 (0.56, 0.82) |
| Prior infection & vaccination: last event >6 months ago | 0.91 (0.78, 1.06) | 0.94 (0.81, 1.10) | 0.88 (0.76, 1.03) |
| Unknown | 1.01 (0.79, 1.29) | 1.05 (0.88, 1.25) | 0.97 (0.80, 1.18) |

## References

1. Polack FP, Thomas SJ, Kitchin N, et al. Safety and Efficacy of the BNT162b2 mRNA Covid-19 Vaccine. New England Journal of Medicine 2020; 383(27): 2603–15.

2. Altarawneh HN, Chemaitelly H, Ayoub HH, et al. Effects of Previous Infection and Vaccination on Symptomatic Omicron Infections. New England Journal of Medicine 2022; 387(1): 21–34.

3. Feikin DR, Higdon MM, Abu-Raddad LJ, et al. Duration of effectiveness of vaccines against SARS-CoV-2 infection and COVID-19 disease: results of a systematic review and meta-regression. The Lancet 2022; 399(10328): 924–44.

4. Qasmieh SA, Robertson MM, Rane MS, et al. The Importance of Incorporating At-Home Testing Into SARS-CoV-2 Point Prevalence Estimates: Findings From a US National Cohort, February 2022. JMIR Public Health Surveillance 2022; 8(12): e38196.

5. Stein C, Nassereldine H, Sorensen RJD, et al. Past SARS-CoV-2 infection protection against re-infection: a systematic review and meta-analysis. The Lancet 2023; 401(10379): 833–42.

6. Sun K, Tempia S, Kleynhans J, et al. Rapidly shifting immunologic landscape and severity of SARS-CoV-2 in the Omicron era in South Africa. Nat Commun 2023; 14(1): 246.

7. Suryawanshi R, Ott M. SARS-CoV-2 hybrid immunity: silver bullet or silver lining? Nature Reviews Immunology 2022; 22(10): 591–2.

8. Stamatatos L, Czartoski J, Wan YH, et al. mRNA vaccination boosts cross-variant neutralizing antibodies elicited by SARS-CoV-2 infection. Science 2021; 372(6549): 1413–8.

9. Bobrovitz N, Ware H, Ma X, et al. Protective effectiveness of previous SARS-CoV-2 infection and hybrid immunity against the omicron variant and severe disease: a systematic review and meta-regression. The Lancet Infectious Diseases 2023; 23(5): 556–67.

10. Chin ET, Leidner D, Lamson L, et al. Protection against Omicron from Vaccination and Previous Infection in a Prison System. New England Journal of Medicine 2022; 387(19): 1770–82.

11. Powell AA, Kirsebom F, Stowe J, Ramsay ME, Lopez-Bernal J, Andrews N, Ladhani SN. Protection against symptomatic infection with delta (B.1.617.2) and omicron (B.1.1.529) BA.1 and BA.2 SARS-CoV-2 variants after previous infection and vaccination in adolescents in England, August, 2021-March, 2022: a national, observational, test-negative, case-control study. The Lancet Infectious Diseases 2023; 23(4): 435–44.

12. de Gier B, Huiberts AJ, Hoeve CE, et al. Effects of COVID-19 vaccination and previous infection on Omicron SARS-CoV-2 infection and relation with serology. Nat Commun 2023; 14(1): 4793.

13. Lewis N, Chambers LC, Chu HT, et al. Effectiveness Associated With Vaccination After COVID-19 Recovery in Preventing Reinfection. JAMA Netw Open 2022; 5(7): e2223917-e.

14. Hall V, Foulkes S, Insalata F, et al. Protection against SARS-CoV-2 after Covid-19 Vaccination and Previous Infection. New England Journal of Medicine 2022; 386(13): 1207–20.

15. Abu-Raddad LJ, Chemaitelly H, Ayoub HH, et al. Effect of mRNA Vaccine Boosters against SARS-CoV-2 Omicron Infection in Qatar. New England Journal of Medicine 2022; 386(19): 1804–16.

16. Lind ML, Dorion M, Houde AJ, et al. Evidence of leaky protection following COVID-19 vaccination and SARS-CoV-2 infection in an incarcerated population. Nature Communications 2023; 14(1): 5055.

17. Bi Q, Lessler J, Eckerle I, et al. Insights into household transmission of SARS-CoV-2 from a population-based serological survey. Nat Commun 2021; 12(1): 3643.

18. McLean HQ, Grijalva CG, Hanson KE, et al. Household Transmission and Clinical Features of SARS-CoV-2 Infections. Pediatrics 2022; 149(3).

19. Centers for Disease Control and Prevention. COVID-19 ACIP Vaccine Recommendations. 2023. https://www.cdc.gov/vaccines/hcp/acip-recs/vacc-specific/covid-19.html (accessed June 20 2023).

20. Jones JM, Manrique IM, Stone MS, et al. Estimates of SARS-CoV-2 Seroprevalence and Incidence of Primary SARS-CoV-2 Infections Among Blood Donors, by COVID-19 Vaccination Status - United States, April 2021-September 2022. Morbidity and Mortality Weekly Report 2023; 72(22): 601–5.

21. Centers for Disease Control and Prevention. Isolation and Precautions for People with COVID-19. 2023. https://www.cdc.gov/coronavirus/2019-ncov/your-health/isolation.html (accessed June 20 2023).

22. Chia WN, Zhu F, Ong SWX, et al. Dynamics of SARS-CoV-2 neutralising antibody responses and duration of immunity: a longitudinal study. The Lancet Microbe 2021; 2(6): e240–e9.

